# A Machine Learning Approach to Predict Cognitive Decline in Alzheimer’s Disease Clinical Trials

**DOI:** 10.1101/2024.09.03.24312481

**Authors:** Bhargav T. Nallapu, Kellen K. Petersen, Tianchen Qian, Idris Demirsoy, Elham Ghanbarian, Christos Davatzikos, Richard B. Lipton, Ali Ezzati, Alzheimer’s Disease Neuroimaging Initiative

## Abstract

**Background:** Of persons randomized to the placebo arm of Alzheimer’s Disease (AD) treatment trials, 40% do not show cognitive decline over 80 weeks of follow-up. Identifying and excluding these individuals from both arms of randomized clinical trials (RCTs) of AD has the potential to increase power to detect treatment effects.

**Objectives:** We aimed to develop machine learning-based predictive models to identify persons unlikely to show decline on placebo treatment over 80 weeks.

**Method:** We used the data from 1072 patients with mild dementia and biomarker evidence of amyloid burden from the placebo arm of EXPEDITION3 trial. Participants were identified as those who demonstrated Clinically Meaningful Cognitive Decline (CMCD, change in ADAS-Cog≥3) or Cognitive Stable (CS, change in ADAS-Cog<3) at final visit of the trial (week 80). Machine learning-based classifiers were trained to classify participants into CMCD vs. CS groups using combinations of demographics, neuropsychological tests (NP) and biomarkers, including APOE4 genotype and volumetric MRI. The results were developed in 70% of the EXPEDITION3 placebo sample (EXP_train_) using 5-fold cross-validation. Trained models were then used to classify the participants in an internal validation sample (EXP_valid)_ and an external matched sample from the Alzheimer’s Disease Neuroimaging Initiative (ADNI) study.

**Result:** Participants selected from the EXPEDITION3 trial were on average 72.7(±7.7) years old, 59% were female. CMCD was observed in 55.8% of participants of EXPEDITION3 at final visit. In the independent validation sample within the EXPEDITION3 data, all the models showed high sensitivity and modest specificity. Positive predictive values (PPVs) of models were at least 11% higher than base prevalence of CMCD observed at the end of the trial. The subset of matched ADNI participants (ADNI_AD_) were on average 74.5(±6.4) years old and 46% female. The models that were validated in ADNI_AD_ also showed high sensitivity, modest specificity and PPVs of at least 15% higher than the base prevalence in ADNI_AD._

**Conclusion:** Our results indicate that predictive models have the potential to improve the design of AD trials through selective inclusion criteria for participants expected to decline and exclusion of those expected to remain stable.

## 1 Introduction

The promise of Disease-Modifying Therapies (DMTs) for Alzheimer’s Disease (AD) lies in their potential to delay or slow the clinical progression of by addressing disease pathologies before they reach a stage of irreversible cell death. These therapies aim to intervene in the underlying mechanisms of the disease, potentially altering their course and providing more effective treatment outcomes ^1,2^. Therefore, the primary outcome in randomized clinical trials (RCTs) of AD typically involves assessing changes in clinical and cognitive outcomes. One of the objectives of these trials is to show directional concordance by correlating the deceleration of cognitive and functional decline with alterations in biomarkers that index the core pathologies.

Most clinical trials in AD have not been successful ^3,4^. Clinical trials fail for several reasons. Biological variability of the disease, such as the phenotypic heterogeneity in case of AD, can obscure the effects of a treatment ^5,6^. Biological heterogeneity in AD, the role of concomitant pathologies and comorbidities likely leads to variability in participant’s response to treatment. Many trials address this heterogeneity at the time of recruitment by imposing strict inclusion and exclusion criteria including family and personal clinical history, clinical stage of the disease, and in-vivo biomarkers ^7,8^, adding to the complexity and cost of study enrollment^9^. Studies rarely consider the uneven rates of expected cognitive decline among eligible trial participants. Prior work in the placebo arm of randomized trials and in observational studies show that up to half of patients with AD do not show meaningful cognitive decline over the course of 18 to 24 months even among individuals who are amyloid positive ^10–12^. If the study endpoint includes a reduction in the rate of decline on active treatment in comparison with placebo, inclusion of individuals likely to show no cognitive decline could be highly impactful on effect size and power.

To apply this insight to the design of clinical trials requires that we identify and exclude these individuals, who are unlikely to decline, from enrollment. In this study, our primary objective was to use data from the placebo arm of a phase III RCT to develop machine learning (ML) predictive models to identify and exclude individuals anticipated not to show cognitive decline by the end of the trial, based on their baseline characteristics. We used data from the placebo arm data from EXPEDITION3, a phase III RCT of Solanezumab for development and internal validation of models and data from the Alzheimer’s Disease Neuroimaging Initiative (ADNI) for external validation of our findings. It has been shown that individualized predictions from ML models, particularly using imaging markers, can be used to inform sample size calculations or to considerably improve statistical power for detecting treatment effects ^13^. Using similar approaches, we performed plasmode simulations of treatment effects using randomized subsets of EXPEDITION3 placebo arm data informed by our imaging-based predictive models to compare the statistical power with the classical modelling that does not include any predictors. Additionally, we investigated whether incorporating 6-month change in cognition enhanced the models’ performance. We discuss how our findings hold implications for the development of models capable of enriching AD trials, paving the way for more increased success of future trials.

## 2 Methods

### 2.1 Design and Participants of the Studies

We used data from a clinical trial EXPEDITION3 (clinicalTrials.gov number NCT01900665) and the ADNI study whose recruitment was designed to simulate a clinical trial ^14^. EXPEDITION3 was a placebo-controlled phase-3 global clinical trial for Solanezumab, a humanized monoclonal antibody that increases clearance of soluble Aβ from the brain. The trial was conducted by Eli Lilly and Company with a primary objective of decreasing cognitive decline in mild dementia due to AD. The trial was conducted across 198 sites in 11 countries with institutional review board approval at each institution and a written informed consent from all participants. The data used in this study was from the placebo arm of the trial.

For external validation, we use data from *ADNI*, an ongoing cohort with the cycles ADNI-1, ADNI-GO, ADNI-2, and ADNI-3 across numerous participating institutions. Data used in this study were obtained from the ADNI database (adni.loni.usc.edu). The ADNI was launched in 2003 as a public-private partnership, led by Principal Investigator Michael W. Weiner, MD. The primary goal of ADNI has been to test whether serial magnetic resonance imaging (MRI), positron emission tomography (PET), other biological markers, and clinical and neuropsychological assessment can be combined to measure the progression of mild cognitive impairment (MCI) and early Alzheimer’s disease (AD). For up-to-date information, see www.adni-info.org. ADNI was approved by the institutional review boards at all the participating institutions. Written informed consent was obtained by or on behalf of all the participants at each site.

#### 2.1.1 EXPEDITION3 – Placebo Arm Population

EXPEDITION3 trial included participants aged 55 to 90 years old, with mild AD without depression, besides other inclusion and exclusion criteria. Participants who were in need, were allowed to receive therapy - including treatments for symptoms of dementia (acetylcholinesterase inhibitors and memantine) and nondrug treatments to ensure that they continued receiving the standard of care for Alzheimer’s disease. The recruitment and study methods of were described elsewhere ^12^. Mild AD at screening was determined by a score of 16 to 26 in the Mini Mental State Examination (MMSE; score range: 0-30, higher scores indicate better cognition ^15^) accompanied by florbetapir positron emission tomography (PET) scan or cerebrospinal fluid (CSF) result consistent with the presence of amyloid pathology. The absence of depression was defined as a Geriatric Depression Scale (GDS; score range: 0–15, higher scores indicate more severe depression ^16^) score of less than or equal to 6 (on the staff-administered short form). Data from 894 participants who were in the placebo arm of the trial and successfully completed the EXPEDITION3 trial were utilized for this study.

#### 2.1.2 Mild AD without depression in ADNI

ADNI, across all 4 cycles, included participants predominantly in the same age group as EXPEDITION3. ADNI also has most of the neuropsychological instruments that were used in the screening of EXPEDITION3, including MMSE and GDS. The amyloid pathology in brain in ADNI is also measured by similar florbetapir (AV45) PET processing methods used by EXPEDITION3. We selected the subset of ADNI participants (ADNI_AD_) who had scores of MMSE and GDS in the range of inclusion criteria of EXPEDITION3 and those who had amyloid burden determined by amyloid PET or CSF, with the diagnosis of Dementia at baseline. A total of 107 participants from ADNI who had 2 years of follow up data were eligible for this study.

#### 2.1.3 Study Outcomes

The primary outcome measure of EXPEDITION3 trial was change from baseline in Alzheimer’s Disease Assessment Scale (higher scores indicating greater cognitive impairment) ^17^ – a 14 Item Cognitive Subscore ^18^ (ADAS-Cog14; score range, 0–90) whereas the secondary outcomes included the change from baseline in the 11 item score of the same (ADAS-Cog11; score range, 0–70). The cognitive assessments in ADNI included ADAS-Cog11 and a 13-item subscore of ADAS across all the phases and participant visits. We chose ADAS-Cog11 as our primary outcome of interest in this study, as this measure was available across both the EXPEDITION3 and ADNI datasets.

Longitudinal cognitive change was defined by the change in ADAS-Cog11 scores from baseline to the end of trial. The variability of ADAS-cog11 scores across visits can exceed the annual rate of change in trials, which is potentially attributable to measurement error and to genuine variation in cognitive performance from day to day ^19–21^. To minimize the effects of such variations on our model outcome, we calculated the change from baseline at both the final as well as one of the visits leading up to the final visit of the studies. In addition to considering the statistically significant changes in cognition, we wanted to ensure that the change is clinically relevant. Although any decline in cognition is undesirable from a patient’s point of view, a clinically meaningful cognitive decline (CMCD) had been suggested if there is a decline of 3 or more points on ADAS-cog11 ^22^. This will mitigate the impact of minor variations in cognitive performance inherent in ADAS-cog11, the primary measure for cognitive decline, on the model’s performance, consistent with findings from prior research. ^22,23^. Thus, we defined the primary outcome of our study to be whether a participant shows CMCD or not (CMCD: ADAS-Cog11_Week80_ – ADAS-Cog11_baseline_ ≥ 3). In the case of ADNI, there were follow up visits of participants every 3 months, 6 months and annually. We calculated the same outcome in ADNI at the 2 years follow up visit from the baseline visit (CMCD: ADAS-Cog11_2yrs_ – ADAS-Cog11_baseline_ ≥ 3).

### 2.2 Other study measures and features

We designed the study to explore if we can predict the participants who would show cognitive decline by the end of the study duration using the baseline measures. Since the goal of this study is to better inform clinical trials towards enrichment strategies, we try to utilize the maximum information available from the screening visits which includes multiple neuropsychological measures and volumetric imaging measures of several brain regions. The following measures are considered in the current study:

- Demographics (D): age, sex, years of education
- Genetics (A): apolipoprotein E (APOE) ε4 alleles (0, 1, 2)
- Volumetric MRI measures (M): Entorhinal Cortex, Hippocampus, Inferior Parietal, Superior and Middle Temporal Cortices
_-_ Clinical characteristics (NP): GDS, Clinical Dementia Rating Sum of Boxes (CDR-SB)_24_
- Cognitive measures (NP): ADAS-Cog11, MMSE
- Functional measures (NP): Alzheimer’s Disease Cooperative Study–Activities of Daily Living (ADCS-ADL) scale (score range, 0–78, with lower scores indicating worse functioning) ^25^, Functional Activity Questionnaire (FAQ) ^26^

Except ADCS-ADL, all the measures were available in both in the EXPEDITION3 trial as well as in ADNI_AD_ dataset.

### 2.3 Predictive models of cognitive stability

#### 2.3.1 Machine Learning models

We used a set of Machine Learning (ML) models to learn the characteristics of both the groups - participants who showed CMCD and those who did not. Each model included some or all of the measures listed in Section 2.2 (D, A, M, NP). ML models are known to be more suitable to handle high-dimensional data where classical multivariate statistical models might be susceptible to noise, especially when the sample size is small ^27^. ML models have also been proven to be effective tools for predictions of outcomes in AD. More details on the benefits of ensemble ML methods, that produce output by combining the low-level, simpler predictive models on subsets of features, are described elsewhere ^28^ ^29^. We chose one ensemble ML model – Random Forest Classifier, and another supervised learning algorithm Linear Discriminant Analysis (LDA) to classify CN and CMCD groups.

The development and validation of the models was done using the scikit-learn libraries in Python (3.9). All the models were assessed in terms of their Sensitivity of classifying actual CMCD (%), Specificity of classifying cognitively stable participants (%), Positive Predictive Value (PPV,%), Negative Predictive Value (NPV,%) within 95% confidence interval (CI) and finally the area under the ROC curve (AUC), all in comparison to the base prevalence of percentage of the population that showed CMCD.

#### 2.3.2 Predictive models of cognitive decline in EXPEDITION3

We developed models that could predict whether a participant in the placebo arm of EXPEDITION3 would show CMCD at the end of Week 80 of the trial. To finetune the ML models, we trained the models using 70% of the data (EXP_train_) using 5-fold cross-validation. Trained models were used to classify the participants in independent sample, the remaining 30% of the data (EXP_valid_). The performances of the models were evaluated using the change in cognition at the end of Week 80 (as described in Section 2.1.3).

##### Plasmode simulations of treatment effects using predictive models

To compare the statistical power of different approaches using our predictive models to a more classical analysis in a clinical trial setting, we conducted plasmode simulations, where hypothetical trial data is generated from the available EXPEDITION3 placebo arm data. First, the available EXPEDITION3 placebo arm data was divided into two halves at random – one for training the predictive models and the other for performing the plasmode simulations (N_pl_). In each of the simulations, the hypothetical trial data consists of random placebo and treatment groups of same sample size drawn from non-overlapping subsets of N_pl_. A classical analysis of treatment effects can be performed using linear regression for a continuous outcome similar to previously described ANOVA-CHANGE models ^30^.

We considered two approaches to incorporate our predictive models in either trial enrollment or post-study analyses and evaluated the added value of our predictive models in terms of increasing the power for detecting treatment effects. (I) Using the predictive models to inform the trial enrollment and (II) Using the individualized prediction of clinical trial outcome from the predictive models as a prognostic variable in post-study analyses.

##### Approach I – Using Predictive Model to Guide Enriched Enrollment

In each simulation, we simulated two hypothetical clinical trials: one with enriched enrollment, whose sample consisted only the subset of participants in N_pl_ that were predicted to show CMCD (N_CMCD_) using their baseline MRI data and our predictive models; the other without enriched enrollment, whose sample of the same size (N_CMCD_) is randomly drawn from N_pl_. Given a desired treatment effect size, a constant treatment effect is added to participants randomized to the treatment arm in both simulated trials. The same random noise was added to both simulated trials to ensure variation across simulations. We used the unadjusted analysis **(Equation 1, Supplementary Method 1)** to test for null effect for both trials. The powers based on the two enrollment strategies were compared for a range of treatment effect sizes.

##### Approach II – Using Predictive Model to Construct Prognostic Variable

In each simulation, we simulated a hypothetical clinical trial with sample randomly drawn from N_pl_, with a constant treatment effect size and random noises added as in Approach I. We considered two analyses to test for null effect: one being the unadjusted analysis **Equation 1 (Supplementary Method 1)**; the other being an adjusted analysis **(Equation 2, Supplementary Method 1)** that incorporates as prognostic variable a predictor that captures the individual’s prognosis of showing CMCD at the end of 2 years, which is obtained by slightly modifying the output of our predictive models. For each analysis, we obtained through simulation the smallest n to achieve 80% power. The smallest sample sizes for the two analyses were compared for a range of treatment effect sizes.

The plasmode simulations were carried out using R studio. A more detailed account of the simulations is available in **Supplementary Method 1** as well as in previously described work ^13^.

#### 2.3.3 Predicting cognitive decline using short-term change in cognition

With the available longitudinal data from follow up visits in both EXPEDITION3 and ADNI_AD_, we calculated the near-term change in cognition (ΔADAS-cog and ΔFAQ) from baseline at Week 28 in the case of EXPEDITION3 and 6-month follow up visit in ADNI_AD_. We trained and validated a new model - D+A+NP+M+Δcog6m, by adding ΔADAS-cog and ΔFAQ to the measures used in the D+A+ NP+M model, in the EXPEDITION3 data, as described in Section 2.3.2. We then assessed this new model with the outcome of CMCD comparing it to the cognition status at the 2 years follow up in ADNI_AD._

#### 2.3.4 External validation of predictive models using ADNI study data

To further validate the robustness of the models in predicting cognitive decline in individuals using baseline characteristics, we used the models trained and finetuned models on the same EXP_train_ subset and assessed the performance on a completely independent dataset ADNI_AD_ evaluated against the change in cognition at the end of the 2 years follow up visit of the participants. **Figure 1** provides an overview of the study design and details of the training and validation procedures.

**Figure 1.**
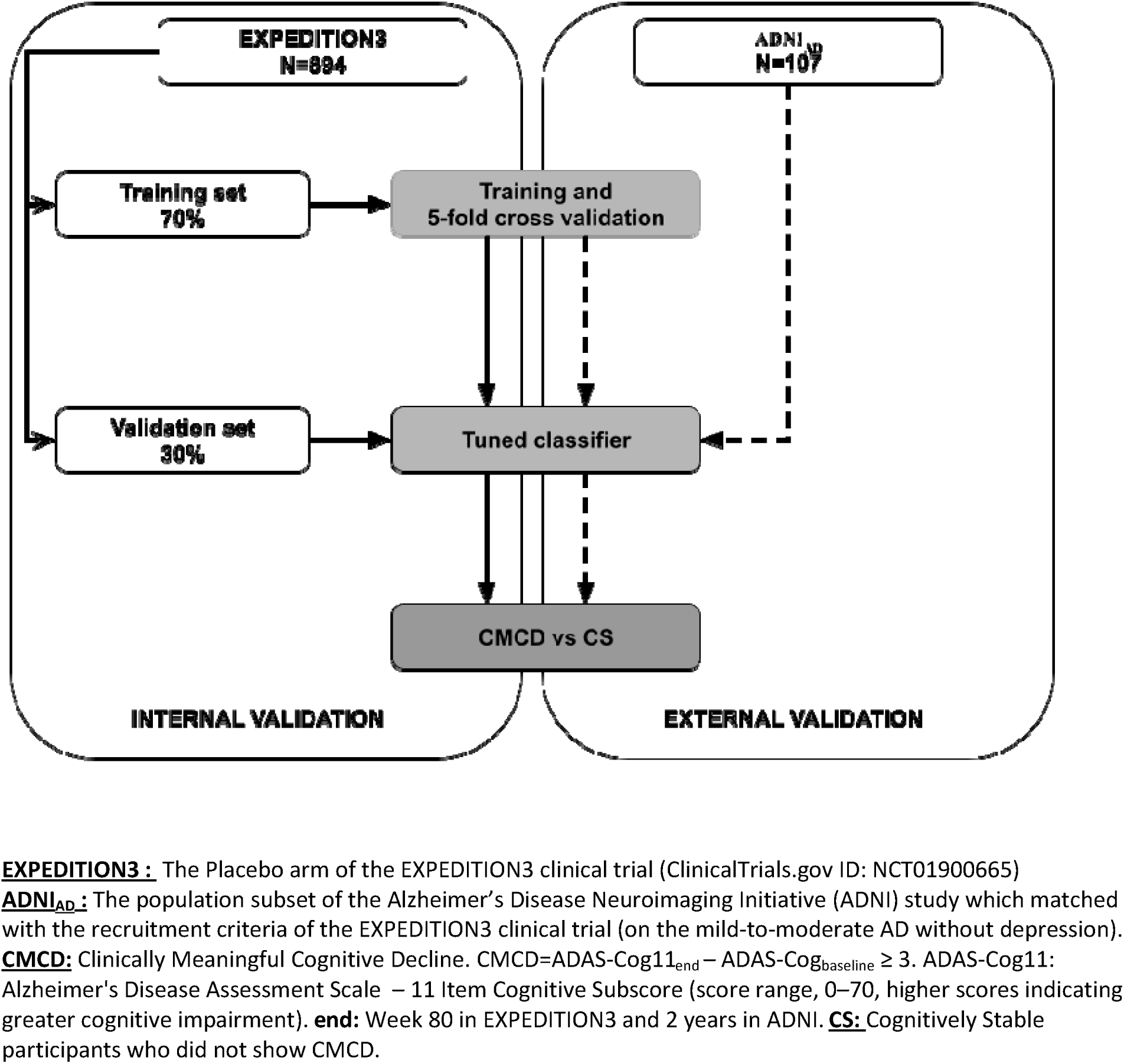
Study Design

### 2.4 Data Availability

Data used in these analyses are from the Eli-Lilly trial: EXPEDITION3 (ClinicalTrials.gov Identifier: NCT01900665). Eli-Lilly makes patient-level data available from Lilly-sponsored studies on marketed drugs for approved uses following approval by regulators in the US and EU and after the primary manuscript describing the results has been accepted for publication, whichever is later. Lilly is one of several companies that provide this access through the website clinicalstudydatarequest.com. Qualified researchers can submit research proposals and request anonymized data to test new hypotheses. Lilly’s data-sharing policies are provided on the clinicalstudydatarequest.com site under the Study Sponsors page.

## 3 Results

### 3.1 Baseline Characteristics

Figure 2A depicts the change in ADAS-cog11 in the placebo population across different weeks into the trial. Within the placebo arm, 498 participants (56%) showed cognitive decline during the 80 weeks of placebo treatment (Figure 2B). The decliner group had a slightly lower average age (71.8±7.8 years) compared to the stable group (73.8±7.3 years). In both the decliner and stable groups, 64% of the participants were carriers of at least one APOE ε4 allele. Of all participants, 59% were female, with 61% of the decliner group and 58% of the stable group being female. The average baseline ADAS-cog11 score for all participants was 18.2(±6.5), with the decliner group having a slightly higher average score (18.8±6.9) compared to that of the stable group (17.6±5.9). The average baseline MMSE (Mini-Mental State Examination) score for all participants was 22.8(±2.9), with the decliner group having a slightly lower average score (22.3±2.9) compared to that of stable group (23.4±2.7). The data from the ADNI_AD_ included 107 participants, with 81% of them being APOE ε4 allele carriers and 48% female. 53% of ADNI subset showed CMCD. In the ADNI_AD_ subset, the average baseline ADAS-cog11 score (15.4±6.6) was lower and the average baseline MMSE score (24.3±1.6) than that of the EXPEDITION3 population. **Table 1** summarizes all the characteristics across EXPEDITION3 and ADNI_AD_ populations.

**Figure 2:**
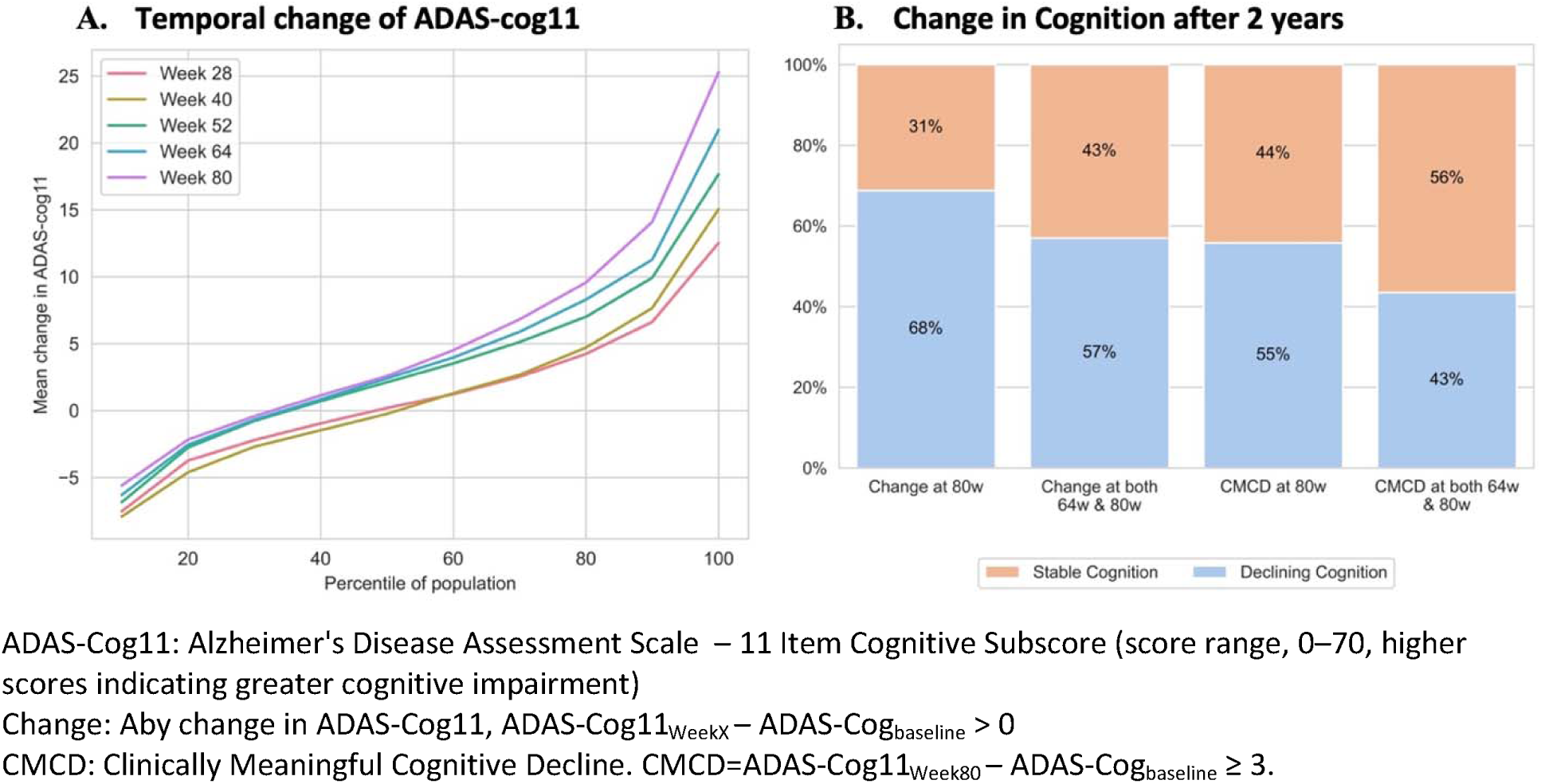
Change in Cognition in EXPEDITION3 placebo arm participants after 2 years

**Table 1.** Participant Characteristics in EXPEDITION3* (placebo arm) and ADNI_AD_ studies.

|  | EXPEDITION3-Placebo Arm |  |  | ADNI*<sub>AD</sub> |  |  |
| --- | --- | --- | --- | --- | --- | --- |
|  | All | Decliner** | Stable | All | Decliner | Stable |
| Count, % | 894 | 498(57) | 392(43) | 107 | 57(53%) | 50(47%) |
| Age, mean (SD), y | 72.7(7.7) | 71.8(7.8) | 73.8(7.3) | 73.5(7.4) | 72.1(8.2) | 75.1(6.1) |
| Female, % | 59% | 61% | 58% | 48% | 43% | 54% |
| Education, mean (SD), y | 13.6(3.7) | 13.5(3.9) | 13.8(3.5) | 15.2(2.6) | 15.2(2.4) | 15.1(2.9) |
| ADAS-cog11 | 18.2(6.5) | 18.8(6.9) | 17.6(5.9) | 15.4(6.6) | 17.4(7.0) | 13.1(5.2) |
| MMSE | 22.8(2.9) | 22.3(2.9) | 23.4(2.7) | 24.3(1.6) | 23.8(1.9) | 24.8(1.2) |
| ADL | 67.3(8.7) | 66.7(9.1) | 68.0(8.3) |  |  |  |
| FAQ | 10.1(6.9) | 10.7(7.0) | 9.4(6.8) | 6.2(7.2) | 7.7(6.5) | 4.5(7.6) |
| GDS | 1.6(1.5) | 1.7(1.5) | 1.6(1.5) | 1.2(1.2) | 1.0(1.1) | 1.4(1.3) |
| CDR | 3.8(1.8) | 4.0(1.9) | 3.5(1.8) | 2.4(2.3) | 3.2(2.5) | 1.4(1.5) |
| APOE ε4 allele(s) |  |  |  |  |  |  |
| 0 | 287(32) | 161(32) | 126(31) | 20(18) | 11(19) | 9(18) |
| 1 | 459(51) | 256(51) | 203(51) | 70(65) | 37(64) | 33(66) |
| 2 | 124(13) | 67(13) | 57(14) | 17(15) | 9(15) | 8(16) |

**EXPEDITION3:** The placebo arm of the EXPEDITION3 clinical trial. \*- Participants were allowed to continue receiving their therapy including treatments for symptoms of dementia (acetylcholinesterase inhibitors and memantine) and nondrug treatments.
**CDR-SB:** Clinical Dementia Rating Sum of Boxes. **ADAS-Cog11** : Alzheimer's Disease Assessment Scale – 11 Item Cognitive Subscore (score range, 0–70, higher scores indicating greater cognitive impairment). **MMSE:** Mini Mental State Examination (score range: 0-30, higher scores indicate better cognition). **ADL:** Alzheimer's Disease Cooperative Study–Activities of Daily Living scale (ADCS-ADL; score range, 0–78, with lower scores indicating worse functioning). **FAQ:** Functional Activity Questionnaire. **APOE:** Apolipoprotein. **\*ADNI:** Subset of ADNI participants with amyloid-PET SUVR > 1.11, 16 ≤ baseline MMSE score ≤ 26, GDS ≤ 6 (Geriatric Depression Scale, range: 0-15, higher scores indicating more severe depression). \*\* Change from baseline to week 80 in ADAS-cog11, *Stable:* < 3, *Decliner:* ≥3.

### 3.2 Performance of predictive models of cognitive decline

#### Training

The performances of 3 models - D+A+M, D+A+NP and D+A+M+NP – trained with 5-fold cross-validation on EXP_train_(N=574) using the LDA method are summarized in **Table 2**. The D+A+NP model showed a moderate performance across all metrics, with an AUC of 0.63 (±0.04). The D+A+M model had an AUC of 0.71 (±0.04) with a sensitivity of 72.5% (95%CI: 68.8-76.2) and a PPV of 69.3% (95%CI: 65.5-73.1) while the base prevalence (BP) of Decliners was 54.4%. Incorporating both M and NP features in the D+A+NP+M model had similar results as those of the D+A+M model with an AUC of 0.71 (±0.04), a sensitivity of 72.1% (95%CI: 68.4-75.8 and a PPV of 67.8% (95%CI: 64.0-71.6). All the models, when evaluated using the RF method, showed similar or lower performances compared to those with the LDA method (Supplementary Table 1).

**Table 2:** Performances of models classifying CS and CMCD groups in EXPEDITION3 training set.

| N | Base prevalence (%) | Method | Model | Sensitivity, %<br>(95%CI) | Specificity, %<br>(95%CI) | PPV, %<br>(95%CI) | NPV, %<br>(95%CI) | AUC |
| --- | --- | --- | --- | --- | --- | --- | --- | --- |
| Training (70% of the EXPEDITION3 SAMPLE) |  |  |  |  |  |  |  |  |
| 574 | 54.4 | LDA | D+A+NP | 69.3 (65.5-73.1) | 48.7 (44.6-52.8) | 61.6 (57.6-65.6) | 57.3 (53.3-61.3) | 0.63 (0.04) |
|  |  |  | D+A+M | 72.5 (68.8-76.2) | 61.4 (57.4-65.4) | 69.3 (65.5-73.1) | 65.6 (61.7-69.5) | 0.71 (0.04) |
|  |  |  | D+A+NP+M | 72.1 (68.4-75.8) | 58.9 (54.9-62.9) | 67.8 (64.0-71.6) | 64.2 (60.3-68.1) | 0.71 (0.04) |
| Internal Validation (30% of the EXPEDITION3 SAMPLE) |  |  |  |  |  |  |  |  |
| 246 | 56.5 | LDA | D+A+NP | 68.3 (62.5-74.1) | 45.8 (39.6-52.0) | 62.1 (56.0-68.2) | 52.7 (46.5-58.9) | 0.57 (0.03) |
|  |  |  | D+A+M | 68.3 (62.5-74.1) | 50.5 (44.3-56.7) | 64.2 (58.2-70.2) | 55.1 (48.9-61.3) | 0.59 (0.03) |
|  |  |  | D+A+NP+M | 69.1 (63.3-74.9) | 52.3 (46.1-58.5) | 65.3 (59.4-71.2) | 56.6 (50.4-62.8) | 0.61 (0.03) |

**CS:** Cognitively Stable. **CMCD:** Clinically Meaningful Cognitive Decline. <sup>a</sup>70% of the entire dataset is used for training with 5-fold cross validation. <sup>b</sup>30% of the entire dataset, reserved for validating the trained models.
**D:** Demographics (age, sex, years of education). **A:** Apolipoprotein E (APOE) ε4 alleles (0, 1, 2). **M:** Volumetric MRI measures (Entorhinal Cortex, Hippocampus, Inferior Parietal, Superior and Middle Temporal Cortices).
**NP:** Clinical characteristics; Clinical Dementia Rating Sum of Boxes (CDR-SB), Alzheimer's Disease Assessment Scale – 11 Item Cognitive Subscore (ADAS-Cog11; score range, 0–70, higher scores indicating greater cognitive impairment), Mini Mental State Examination (MMSE; score range: 0-30, higher scores indicate better cognition), Alzheimer's Disease Cooperative Study–Activities of Daily Living scale (ADCS-ADL; score range, 0–78, with lower scores indicating worse functioning), Functional Activity Questionnaire (FAQ).
**LDA:** Linear Discriminant Analysis.

#### Internal Validation

We assessed the performance of these trained models on the independent dataset EXP_valid_ (N=246, see **Table 2**). The D+A+NP+M model, which combined baseline neuropsychological and volumetric MRI measures, had better overall performance with an AUC of 0.61 (±0.03), better than the D+A+NP (AUC of 0.57±0.03) and D+A+M (AUC of 0.59±0.03) models. The D+A+NP+M model also had better performances across all the other metrics compared to those of the D+A+NP and D+A+M models with a sensitivity of 69.1% (95%CI: 63.3-74.9) and a PPV of 65.3% (95%CI: 59.4-71.2, BP: 56.5%). The models when using the RF method, had slightly lower performances individually, with the model D+A+NP+M performing better than the individual D+A+NP and D+A+M models (Supplementary Table 2).

#### Predicting CMCD using short-term change in cognition

We trained a new model on EXP_train_ using the LDA method, D+A+NP+M+Δcog6m - by combining the ΔADAS-cog and ΔFAQ together with the D+A+M+NP model, the model showed the highest performance (see **Table 3**), an AUC of 0.83 (±0.04), a sensitivity of 74.5% (95%CI: 70.9-78.1) and a PPV of 77.1% (95%CI: 73.7-80.5). When validated on EXP_valid,_ the D+A+NP+M+Δcog6m model again showed the higher performance across all the metrics than the rest of the models with an AUC of 0.74 (±0.03), a sensitivity of 73.4% (95%CI: 67.9-78.9), and a PPV of 79.1% (95%CI: 74.0-84.2, BP: 56.5%). A similar improvement of performance was seen using the RF method, by the addition of Δcog6m to the D+A+NP+M model (Supplementary Table 3).

**Table 3:**
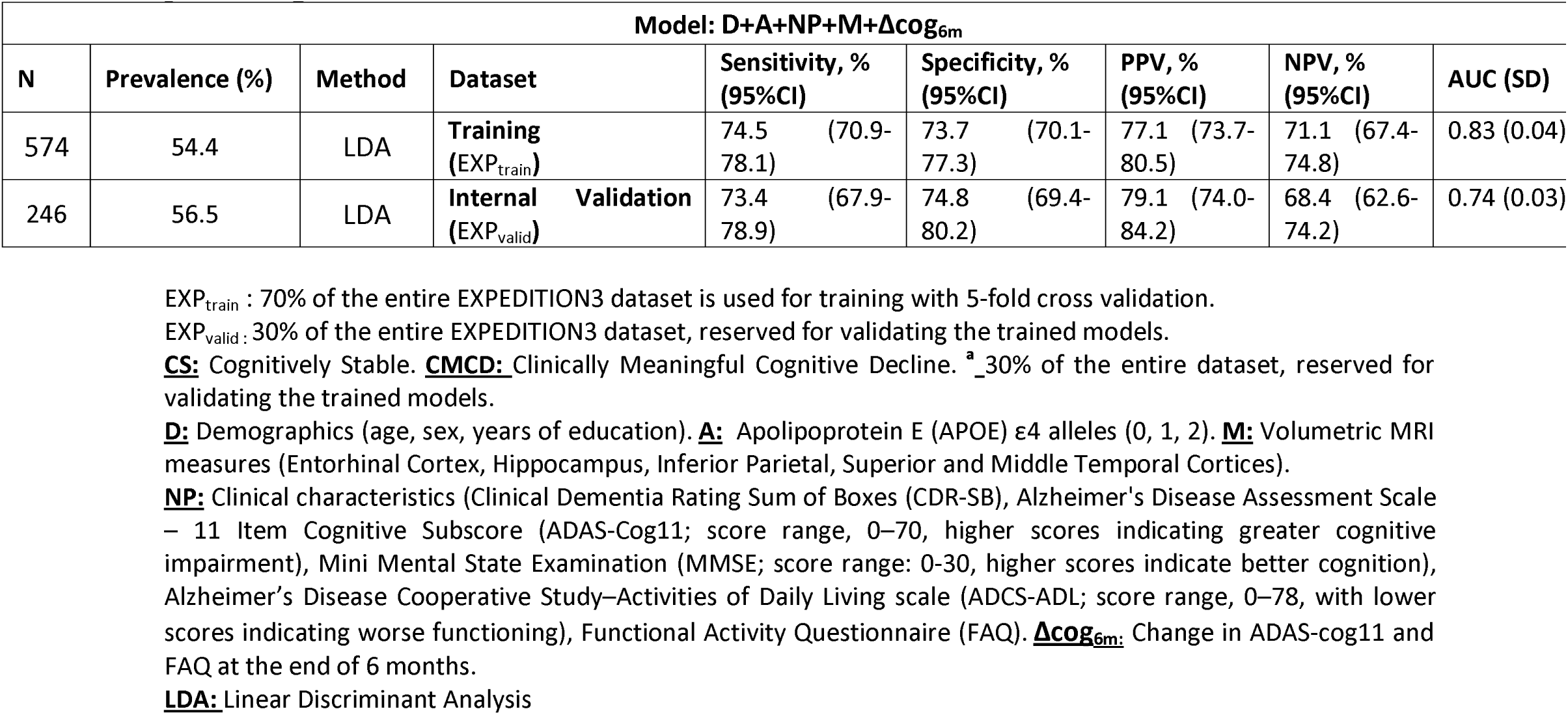
Performance of model including short-term change in cognition over 6 months in predicting CMCD.

| Model: D+A+NP+M+$\Delta\text{cog}_{6m}$ | | | | | | | | |
| --- | --- | --- | --- | --- | --- | --- | --- | --- |
| N | Prevalence (%) | Method | Dataset | Sensitivity, %<br>(95%CI) | Specificity, %<br>(95%CI) | PPV, %<br>(95%CI) | NPV, %<br>(95%CI) | AUC (SD) |
| 574 | 54.4 | LDA | Training<br>(EXP<sub>train</sub>) | 74.5 (70.9-78.1) | 73.7 (70.1-77.3) | 77.1 (73.7-80.5) | 71.1 (67.4-74.8) | 0.83 (0.04) |
| 246 | 56.5 | LDA | Internal Validation<br>(EXP<sub>valid</sub>) | 73.4 (67.9-78.9) | 74.8 (69.4-80.2) | 79.1 (74.0-84.2) | 68.4 (62.6-74.2) | 0.74 (0.03) |

EXP<sub>train</sub> : 70% of the entire EXPEDITION3 dataset is used for training with 5-fold cross validation.
EXP<sub>valid</sub> : 30% of the entire EXPEDITION3 dataset, reserved for validating the trained models.
**CS:** Cognitively Stable. **CMCD:** Clinically Meaningful Cognitive Decline. <sup>a</sup> 30% of the entire dataset, reserved for validating the trained models.
**D:** Demographics (age, sex, years of education). **A:** Apolipoprotein E (APOE) ε4 alleles (0, 1, 2). **M:** Volumetric MRI measures (Entorhinal Cortex, Hippocampus, Inferior Parietal, Superior and Middle Temporal Cortices).
**NP:** Clinical characteristics (Clinical Dementia Rating Sum of Boxes (CDR-SB), Alzheimer's Disease Assessment Scale – 11 Item Cognitive Subscore (ADAS-Cog11; score range, 0–70, higher scores indicating greater cognitive impairment), Mini Mental State Examination (MMSE; score range: 0-30, higher scores indicate better cognition), Alzheimer's Disease Cooperative Study–Activities of Daily Living scale (ADCS-ADL; score range, 0–78, with lower scores indicating worse functioning), Functional Activity Questionnaire (FAQ). **$\Delta\text{cog}_{6m}$ :** Change in ADAS-cog11 and FAQ at the end of 6 months.
**LDA:** Linear Discriminant Analysis

#### External Validation in ADNI_AD_

We then validated the two best performing models - D+A+M+NP and D+A+NP+M+Δcog6m - trained and fine-tuned on the EXP_train_ dataset, on the ADNI_AD_ dataset (see **Table 4**). Both the models using the LDA method were assessed in predicting whether the participants would show CMCD at the end of 2 year follow up in ADNI. The D+A+NP+M model showed a moderately high AUC of 0.77 (±0.04) with a high sensitivity of 87.3% (95%CI: 80.9-93.7), a high NPV of 81.6% (95%CI: 74.2-89.0) and a PPV of 71.6% (95%CI: 63.0-80.2, BP: 52.4%). The D+A+NP+M+Δcog6m had the highest performance among all the models with a high AUC of 0.80 (±0.04) with a sensitivity of 83.6% (95%CI: 76.5-90.7), a high NPV of 80.9% (95%CI: 73.4-88.4) and a high PPV of 79.3% (95%CI: 71.6-87.0, BP: 52.4%).

**Table 4:** Performance of models in predicting CMCD in ADNI_AD_ sample.

| External Validation in ADNI<sub>AD</sub> dataset |  |  |  |  |  |  |  |  |  |  |  |  |
| --- | --- | --- | --- | --- | --- | --- | --- | --- | --- | --- | --- | --- |
| N | Prevalence (%) | Method | Model | Sensitivity, % (95%CI) |  | Specificity, % (95%CI) |  | PPV, % (95%CI) |  | NPV, % (95%CI) |  | AUC (SD) |
| 105 | 52.4 | LDA | D+A+NP+M | 87.3 | (80.9-93.7) | 62.0 | (52.7-71.3) | 71.6 | (63.0-80.2) | 81.6 | (74.2-89.0) | 0.75 (0.04) |
| 105 | 52.4 | LDA | D+A+NP+M+<br>$\Delta\text{cog}_{6m}$ | 83.6 | (76.5-90.7) | 76.0 | (67.8-84.2) | 79.3 | (71.6-87.0) | 80.9 | (73.4-88.4) | 0.80 (0.04) |

**CS:** Cognitively Stable. **CMCD:** Clinically Meaningful Cognitive Decline. <sup>a</sup> ADNI<sub>AD</sub> sample is only used for validating the models that were trained on the EXPEDITION training subset.
**D:** Demographics (age, sex, years of education). **A:** Apolipoprotein E (APOE) $\epsilon 4$ alleles (0, 1, 2). **M:** Volumetric MRI measures (Entorhinal Cortex, Hippocampus, Inferior Parietal, Superior and Middle Temporal Cortices).
**NP:** Clinical characteristics (Clinical Dementia Rating Sum of Boxes (CDR-SB), Alzheimer's Disease Assessment Scale – 11 Item Cognitive Subscore (ADAS-Cog11; score range, 0–70, higher scores indicating greater cognitive impairment), Mini Mental State Examination (MMSE; score range: 0-30, higher scores indicate better cognition), Alzheimer's Disease Cooperative Study–Activities of Daily Living scale (ADCS-ADL; score range, 0–78, with lower scores indicating worse functioning), Functional Activity Questionnaire (FAQ). **$\Delta\text{cog}_{6m}$ :** Change in ADAS-cog11 and FAQ at the end of 6 months.
**LDA:** Linear Discriminant Analysis.

The RF method with the same models showed a slightly better performance for the D+A+NP+M model compared to that of using the LDA method (AUC of 0.77±0.04) but a lower performance for the D+A+NP+M+Δcog6m model (AUC of 0.70±0.03). Detailed results of the validation of the two models on the ADNI_AD_ dataset can be found in Supplementary Table 4.

#### Statistical Power Analysis in Simulated Clinical Trials

Using plasmode simulations, we evaluated the added value of the predictive models of cognitive decline in terms of increased power and reduced sample size, via two approaches to incorporate the predictive models: (I) informing enriched enrollment and (II) constructing prognostic variables for post-study analysis. In (I), for treatment effect sizes ranging from 0.3 to 0.5, the hypothetical trial with enriched enrollment informed by our predictive models yields higher power than the trial without enriched enrollment. The power difference was greater at an effect size of 0.3 (∼83% with enriched enrollment vs ∼50% without). The difference in power reduced for larger effect sizes with the enriched enrollment still achieving higher power (Figure 3A). In (II), the analysis that adjusts for the prognostic variable constructed using our predictive model consistently reduced the minimum required sample size n for 80% power compared to the unadjusted analysis. For example, with an effect size of 0.3, the minimum required sample size was 292 for the adjusted analysis and 336 for the unadjusted analysis. As the effect size increased, sample size requirements decreased for both analyses and the difference between the two analyses also decreased (Figure 3B).

**Figure 3.**
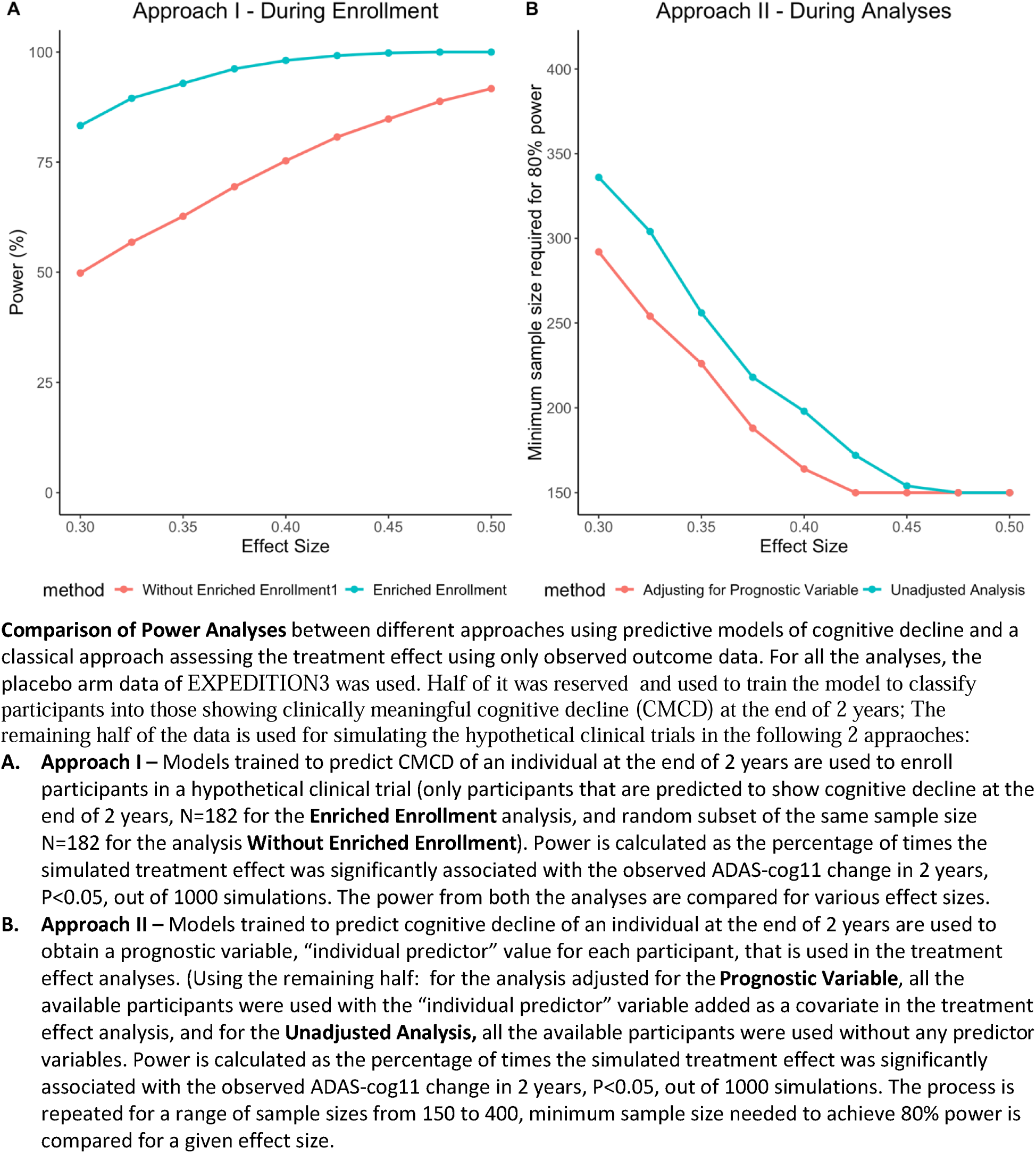
Statistical power analyses of using predictive model in simulating treatment effects in a clinical trial

## 4 Discussion

In this study, we developed models that use baseline characteristics of participants in a clinical trial to classify them into two groups: those who show longitudinal cognitive decline during the trial and those expected to remain cognitively stable. We showed that these models can predict CMCD in 2 years in an internal validation sample and in an independent dataset from the cohort study ADNI whose recruitment was designed to mimic that of a clinical trial. Models using LDA method had better performances than those using RF method in predicting CMCD in both EXP_valid_ and ADNI_AD_ subsets while both the models showed the potential of machine learning models in predicting CMCD. With all the baseline characteristics as features and clinically meaningful cognitive decline (CMCD) as the outcome based on the longitudinal change in ADAS-Cog11 score, predictive models in the independent EXPEDITION3 validation subset (EXP_valid_) had a mean PPV of 65.3%, which was 9% higher than the prevalence of CMCD in the EXP_valid_ subset at the end of the trial. This could represent a considerable degree of enrichment of the decliner group. The same model when tested on ADNI_AD_ subset, had a mean PPV of 71.6%, which is 19% higher than the prevalence in ADNI_AD_ observed at the 2yr follow up visit.

Furthermore, we showed that augmenting the models using baseline characteristics with short-term change in cognition drastically improved the performance of the models in classifying CS participants from CMCD within EXPEDITION3 as well as in the independent dataset ADNI_AD_. Our results highlight the additional value of 6-month change in cognition in predicting the eventual cognitive decline. The addition of 6-month changes in ADAS-cog11 and FAQ scores (Δcog6m) to the model with all the baseline characteristics showed a significantly higher performance in both the validation sample of EXPEDITION3 (EXP_valid_) as well as in the independent ADNI_AD_ sample. The D+A+NP+M+Δcog6m model showed a mean PPV 22.5% higher than the base prevalence of CMCD in the EXP_valid_ sample, with a moderate AUC of 0.73±0.03. The same model performed even better in the independent ADNI_AD_ subset with a mean PPV 27% higher than the prevalence of CMCD at the end 2 year follow up in ADNI, with an AUC as high as 0.80±0.04. We have also shown that our predictive models using imaging biomarkers can be used in both informing enriched trial enrollment and enhancing post-study analysis to increase statistical power and reducing sample size, in line with previous simulation studies ^13^. Our results highlight novel approaches to optimizing the recruitment of RCTs targeting cognitive decline.

The utility of machine learning models in predicting clinically relevant change in cognition in different stages of AD has been demonstrated in various settings ^31,32^. We used 2 different ML methods that offer unique advantages in handling complex AD data. LDA, a supervised classification algorithm, effectively preserves differentiating information across outcomes and reduces input feature dimensions. It is also particularly advantageous for small sample sizes as observed in our independent validation dataset ADNI_AD_. On the other hand, application of a family of ML methods known as ensemble learning has been shown to be highly effective method in predicting clinical outcomes ^28^. RF, an ensemble ML algorithm, was the other method we employed, owing to its resistance to overfitting the training data and its inbuilt feature importance. Both models demonstrated strong performance in predicting cognitive change, suggesting potential for further sophisticated analyses and robust feature selection in future iterations of this study. In the context of clinical trial design, establishing the utility of different clinical and biomarker measures measured at screening or baseline visit may have implications for enrichment strategies with direct impact on costs and success of the trials.

Our work highlights the use of data from the placebo arms of clinical trials in building models that can inform various ADRD studies in future. Over the last 2 decades, only a few of clinical trials targeting AD pathology in brain with an outcome of slowing down cognitive decline succeeded in their goals ^10,33,34^ with the majority of trials failing to meet their outcomes ^35,36^, including the one used in this current study ^12^. Optimizing trial design involves incorporating sequential and adaptive, and enrichment strategies. One approach to enrich AD trials is to include suitable individuals – who are more likely to benefit from therapeutic intervention and exclude those expected to remain cognitively stable and unlikely to show benefit during the limited timeframe of trial. This not only improves the chances of success but also can reduce the costs. Our study demonstrates that predictive models developed using the data from failed trials can have a significant impact on the design of future trials, primarily by enriching the participant recruitment. Another potential application of these models is in conducting post-hoc analyses of completed trials, enabling us to assess the effects of investigational drugs specifically in individuals expected to experience cognitive decline within the trial period.

Several studies have developed models to predict cognitive trajectories in different stages of AD using either longitudinal data from prospective cohorts of ADRD ^28,31^ or the baseline characteristics of a clinical trial population with mild AD ^32^. This study is the first to show models built using both baseline and short-term follow up data are reliable and effective in predicting clinical outcomes. However, our study has some limitations. Firstly, the models in our study used relatively small set of features from volumetric MRI and a small set of neuropsychological scores from each of clinical, cognitive, and functional assessments. Baseline neuropsychological measures are a cross-sectional representation of individual’s cognition, and are prone to measurement errors and high variability ^37^. Another limitation of our models is lack of AD-specific biomarkers such as CSF or plasma amyloid and tau biomarkers. Overall, the performance of models is expected to improve if more detailed, informative data is available. Furthermore, to validate our models, we used a subset from a cohort study whose characteristics were overlapping with the recruitment of the clinical trial, but ADNI should not be considered a trial, and results should be further validated in independent samples from other trials. Finally, we described cognitive decline as opposed to stability in a unidimensional way, whereas it is expected that further classification of decliner populations into rapid vs slow decliners improves the efficacy and applicability of the models.

Notwithstanding the limitations described above, the results of this study from the placebo arm of one AD clinical trial and its validation in an independent population, show a great promise of how predictive models can impact the design of future AD trials. This work can be extended to a more generalizable framework, which exploits the data from placebo groups of the multiple failed trials, providing clinically relevant tools for clinical trial recruitment. Furthermore, in conjunction with treatment data from the trials, this work opens avenues for robust and extensive post-hoc analyses of the treatment effects of DMTs in AD.

## Data Availability

All data produced in the present study are available upon reasonable request to the authors

## Supplementary Material

**Supplementary Figure 1.**
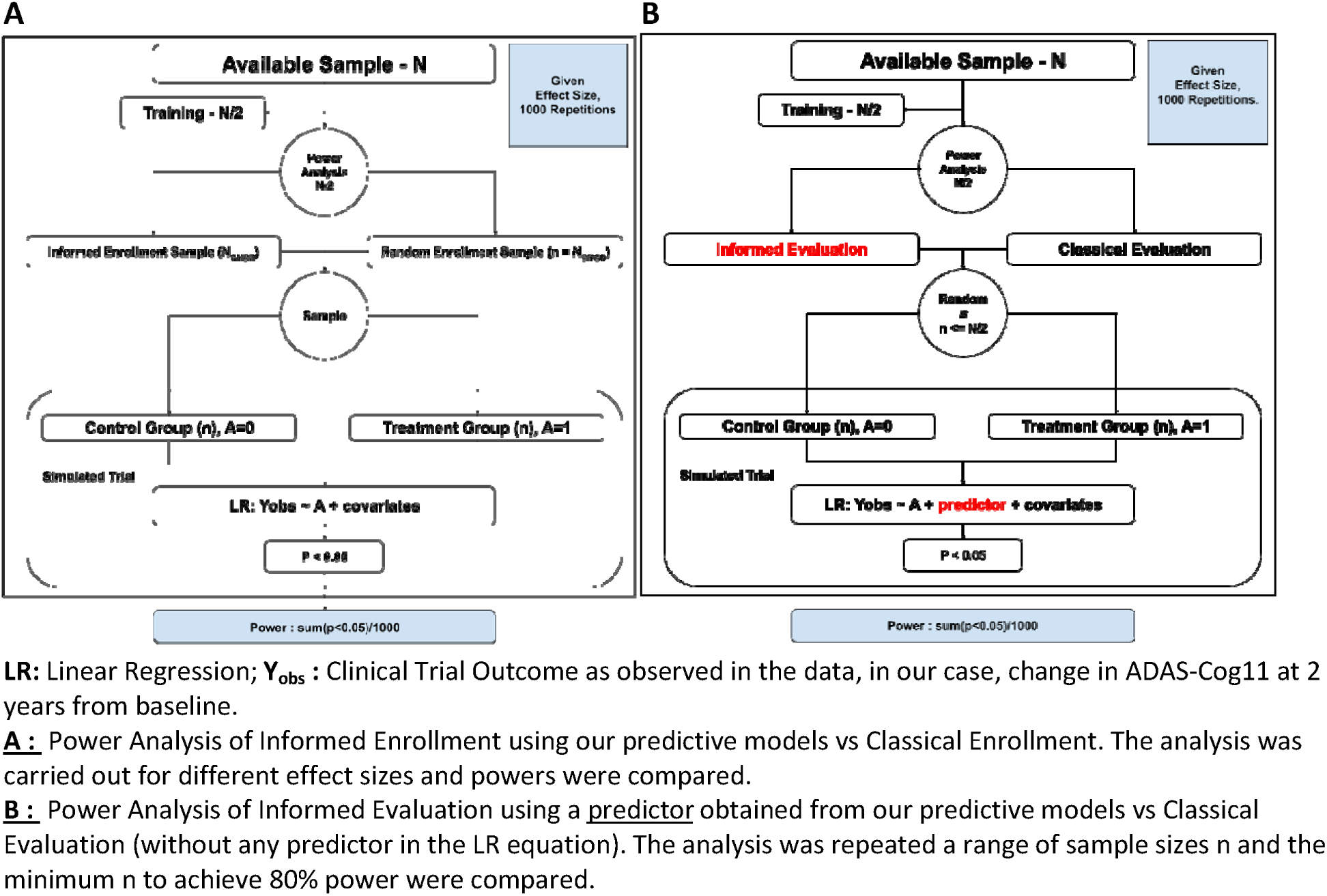
Informed Enrollment in Clinical Trials – Power Analysis

**Supplementary Table 1:**
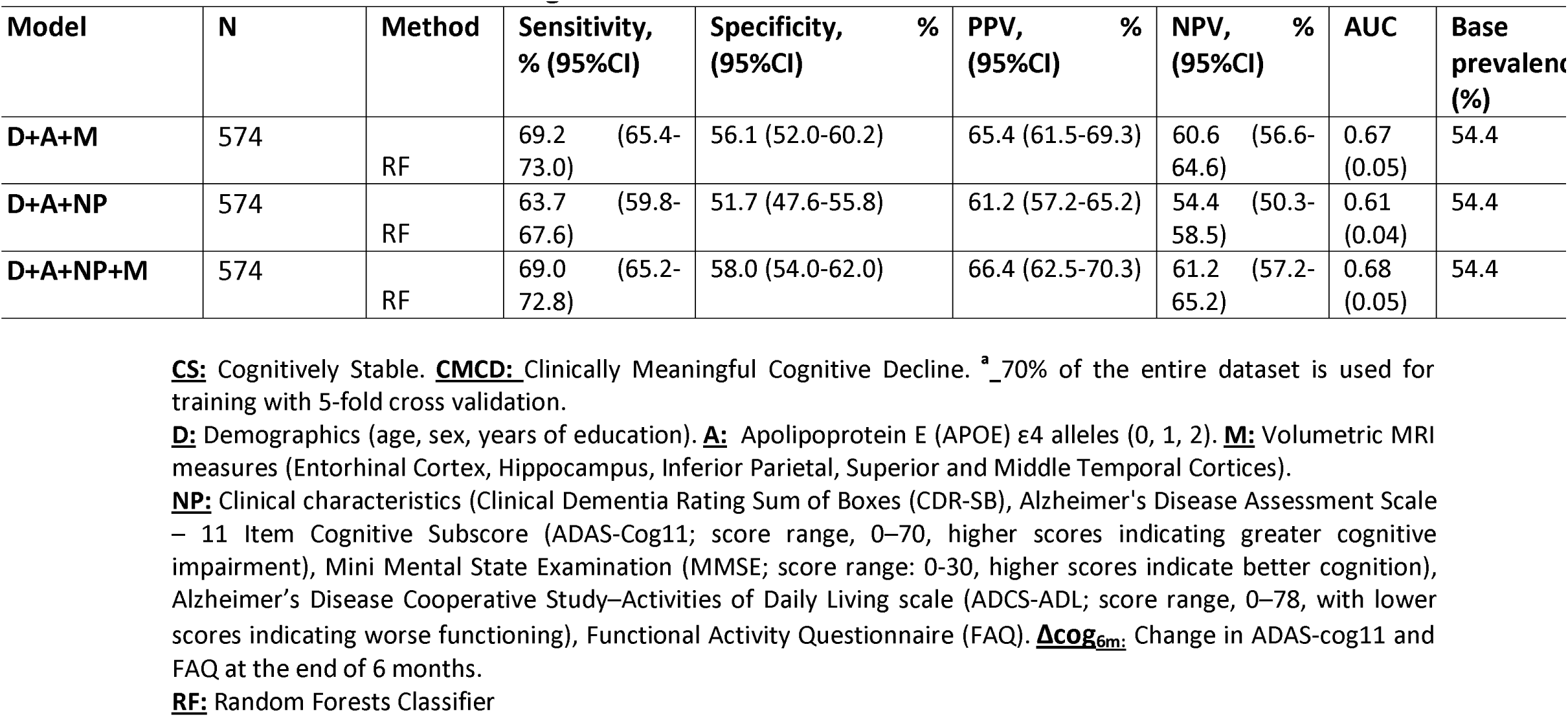
Performances of models classifying CS and CMCD groups in EXPEDITION3 training set.

**Supplementary Table 2:** Performance of models predicting CMCD in EXPEDITION3 validation set.

| Model | N | Method | Sensitivity, %<br>(95%CI) | Specificity, %<br>(95%CI) | PPV, %<br>(95%CI) | NPV, %<br>(95%CI) | AUC (SD) | Prevalence (%) |
| --- | --- | --- | --- | --- | --- | --- | --- | --- |
| **D+A+M** | 246 | RF | 67.6 (61.8-73.4) | 48.6 (42.4-54.8) | 63.1 (57.1-69.1) | 53.6 (47.4-59.8) | 0.58 (0.03) | 56.5 |
| **D+A+NP** | 246 | RF | 64.0 (58.0-70.0) | 50.5 (44.3-56.7) | 62.7 (56.7-68.7) | 51.9 (45.7-58.1) | 0.57 (0.03) | 56.5 |
| **D+A+M+NP** | 246 | RF | 71.9 (66.3-77.5) | 50.5 (44.3-56.7) | 65.4 (59.5-71.3) | 58.1 (51.9-64.3) | 0.61 (0.03) | 56.5 |

**CS:** Cognitively Stable. **CMCD:** Clinically Meaningful Cognitive Decline. <sup>a</sup> 30% of the entire dataset, reserved for validating the trained models.
**D:** Demographics (age, sex, years of education). **A:** Apolipoprotein E (APOE) ε4 alleles (0, 1, 2). **M:** Volumetric MRI measures (Entorhinal Cortex, Hippocampus, Inferior Parietal, Superior and Middle Temporal Cortices).
**NP:** Clinical characteristics (Clinical Dementia Rating Sum of Boxes (CDR-SB), Alzheimer's Disease Assessment Scale – 11 Item Cognitive Subscore (ADAS-Cog11; score range, 0–70, higher scores indicating greater cognitive impairment), Mini Mental State Examination (MMSE; score range: 0-30, higher scores indicate better cognition), Alzheimer's Disease Cooperative Study–Activities of Daily Living scale (ADCS-ADL; score range, 0–78, with lower scores indicating worse functioning), Functional Activity Questionnaire (FAQ). **Δcog<sub>6m</sub>:** Change in ADAS-cog11 and FAQ at the end of 6 months.
**RF:** Random Forests Classifier

**Supplementary Table 3:** Performance of model including short-term change in cognition in predicting CMCD.

| Model: D+A+NP+M+$\Delta\text{cog}_{6m}$ | | | | | | | | | |
| --- | --- | --- | --- | --- | --- | --- | --- | --- | --- |
| Dataset | N | Method | Sensitivity, %<br>(95%CI) | Specificity, %<br>(95%CI) | PPV, %<br>(95%CI) | NPV, %<br>(95%CI) | AUC (SD) | Prevalence (%) |  |
| **Training**<br>(EXP<sub>train</sub>) | 574 | RF | 77.7 (74.3-81.1) | 68.3 (64.5-72.1) | 74.6 (71.0-78.2) | 72.4 (68.7-76.1) | 0.81 (0.05) | 54.4 |  |
| **Internal Validation**<br>(EXP<sub>valid</sub>) | 246 | RF | 77.7 (72.5-82.9) | 66.4 (60.5-72.3) | 75.0 (69.6-80.4) | 69.6 (63.9-75.3) | 0.72 (0.03) | 56.5 |  |

EXP<sub>train</sub> : 70% of the entire EXPEDITION3 dataset is used for training with 5-fold cross validation.
EXP<sub>valid</sub>: 30% of the entire EXPEDITION3 dataset, reserved for validating the trained models.
**CS:** Cognitively Stable. **CMCD:** Clinically Meaningful Cognitive Decline. <sup>a</sup> 30% of the entire dataset, reserved for validating the trained models.
**D:** Demographics (age, sex, years of education). **A:** Apolipoprotein E (APOE) ε4 alleles (0, 1, 2). **M:** Volumetric MRI measures (Entorhinal Cortex, Hippocampus, Inferior Parietal, Superior and Middle Temporal Cortices).
**NP:** Clinical characteristics (Clinical Dementia Rating Sum of Boxes (CDR-SB), Alzheimer's Disease Assessment Scale – 11 Item Cognitive Subscore (ADAS-Cog11; score range, 0–70, higher scores indicating greater cognitive impairment), Mini Mental State Examination (MMSE; score range: 0-30, higher scores indicate better cognition), Alzheimer's Disease Cooperative Study–Activities of Daily Living scale (ADCS-ADL; score range, 0–78, with lower scores indicating worse functioning), Functional Activity Questionnaire (FAQ). **$\Delta\text{cog}_{6m}$ :** Change in ADAS-cog11 and FAQ at the end of 6 months.
**RF:** Random Forests Classifier

**Supplementary Table 4:** Performance of models in predicting CMCD in ADNI_AD_ sample.

| External Validation in ADNI<sub>AD</sub> dataset |  |  |  |  |  |  |  |  |
| --- | --- | --- | --- | --- | --- | --- | --- | --- |
| Dataset | N | Method | Sensitivity, %<br>(95%CI) | Specificity, %<br>(95%CI) | PPV, %<br>(95%CI) | NPV, %<br>(95%CI) | AUC (SD) | Prevalence (%) |
| **D+A+M+NP** | 105 | RF | 84.2 (77.3-91.1) | 46.0 (36.6-55.4) | 64.0 (54.9-73.1) | 71.9 (63.4-80.4) | 0.65 (0.04) | 52.4 |
| **D+A+NP+M+**<br>**Δcog<sub>6m</sub>** | 105 | RF | 69.1 (60.3-77.9) | 68.0 (59.1-76.9) | 70.4 (61.7-79.1) | 66.7 (57.7-75.7) | 0.68 (0.05) | 52.4 |

**CS:** Cognitively Stable. **CMCD:** Clinically Meaningful Cognitive Decline. <sup>a</sup>ADNI<sub>AD</sub> sample is only used for validating the models that were trained on the EXPEDITION training subset.
**D:** Demographics (age, sex, years of education). **A:** Apolipoprotein E (APOE) ε4 alleles (0, 1, 2). **M:** Volumetric MRI measures (Entorhinal Cortex, Hippocampus, Inferior Parietal, Superior and Middle Temporal Cortices).
**NP:** Clinical characteristics (Clinical Dementia Rating Sum of Boxes (CDR-SB), Alzheimer's Disease Assessment Scale – 11 Item Cognitive Subscore (ADAS-Cog11; score range, 0–70, higher scores indicating greater cognitive impairment), Mini Mental State Examination (MMSE; score range: 0-30, higher scores indicate better cognition), Alzheimer's Disease Cooperative Study–Activities of Daily Living scale (ADCS-ADL; score range, 0–78, with lower scores indicating worse functioning), Functional Activity Questionnaire (FAQ). **Δcog<sub>6m</sub>:** Change in ADAS-cog11 and FAQ at the end of 6 months.
**RF:** Random Forests Classifier

**Supplementary Table 5:**
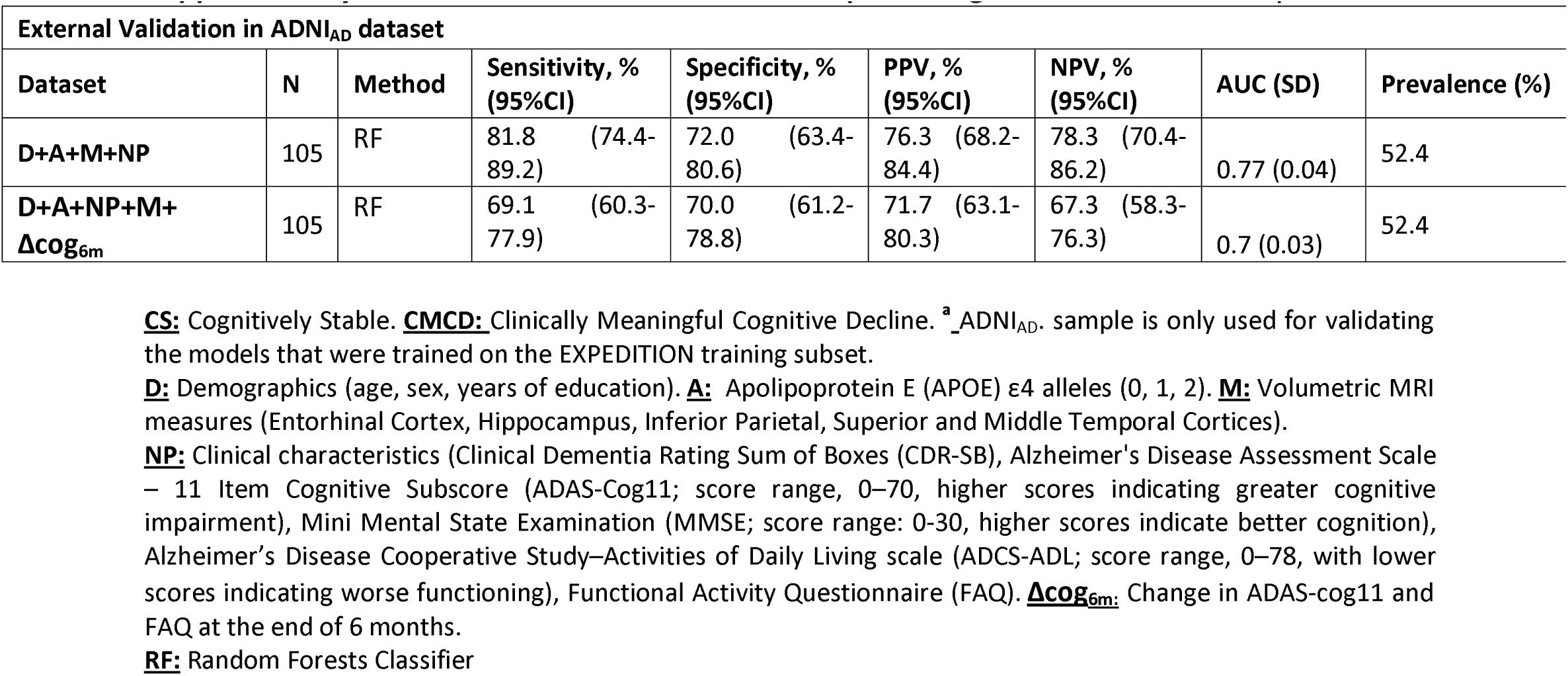
Performance of models in predicting CMCD in ADNI_AD_ sample.

## Supplementary Method 1. Informed Enrollment and Informed Evaluation in Clinical Trials – Statistical Power Analysis

### Plasmode simulations of treatment effects using predictive models

The plasmode simulations were performed in R studio, using core stats packages. To compare the statistical power of different approaches using our predictive models to a more classical analysis in a clinical trial setting, we conducted plasmode simulations, where hypothetical trial data is generated from the available EXPEDITION3 placebo arm data. First, the available EXPEDITION3 placebo arm data was divided into two halves at random – one for training the predictive models and the other for performing the plasmode simulations (N_pl_). In each of the simulations, the hypothetical trial data consists of random placebo and treatment groups of same sample size drawn from non-overlapping subsets of N_pl_. A classical analysis of treatment effects can be performed using linear regression for a continuous outcome (**Equation 1**), similar to previously described ANOVA-CHANGE models ^30^.

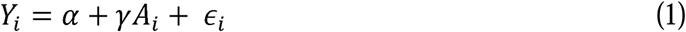

In our study, *Y_i_* represents cognitive decline, defined as the change in ADAS-cog11 score from the baseline observed at 2 years from baseline. *A_i_* represents the treatment indicator, and *ε_i_* represents random error. *γ* is simulated for a range of effect sizes and a is estimated from the data. Within each analysis, 1000 simulations were repeated randomizing the division of treatment and placebo groups. The statistical power of the simulated treatment effect was calculated in both the cases, as the number of times P-value was significant corresponding to the test for treatment effect (*γ*). We explored this for a range of effect sizes, which was defined as *Y_i_* divided by the standard deviation of the outcome *Y_i_*.

We analyzed the performance of our predictive models in the statistical power analysis of treatment effects using two approaches – (I) Using the predictive models to inform the trial enrollment and (II) Using the individualized estimates of clinical trial outcome from the predictive models in treatment effect analyses.

### Approach I – Informed Enrollment

For the analysis of predictive models, the hypothetical clinical trial sample consisted only the subset of participants in N_pl_ that was predicted to show CMCD (N_CMCD_) using their baseline MRI data and our predictive models, an informed enrollment into the clinical trial. For the classical analysis, a random sample of size equal to N_CMCD_ was drawn from N_pl_ as the clinical trial data (**Classical Enrollment**). The analyses for both classical and informed enrollment were carried out using **Equation 1** and their powers were compared for different treatment effect sizes.

### Approach II – Individualized Evaluation

For the analysis of predictive models as well as classical analysis, the hypothetical clinical trial data was randomly drawn from N_pl_. While the statistical power evaluation for classical analysis was carried out using **Equation 1** (**Classical Evaluation**), we used the following equation for evaluating the predictive models, that incorporates a predictor obtained by slightly modifying the output of our models, that represents the individual’s likelihood to show CMCD at the end of 2 years as predicted at baseline (**Individualized Evaluation**).

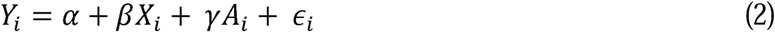

Where *X_i_* represents the predictor obtained from our model and *β* is estimated from the data. The analysis for both classical and individualized evaluation were repeated for a range of sample sizes and the smallest n for which power reached 80% were compared.

## Notes

### Competing Interest Statement

The authors have declared no competing interest.

### Funding Statement

There was no direct funding for this work. The author(s) would like to disclose the following.
Dr. Bhargav T. Nallapu has nothing to disclose; Dr. Kellen K. Petersen has nothing to disclose; Dr. Tianchen Qian has nothing to disclose; Dr. Idris Demirsoy has nothing to disclose; Dr. Elham Ghanbarian has nothing to disclose; Dr. Christos Davatzikos receives research support in part by grants from the National Institute of Health (NIA RF1AG054409); Dr. Richard B. Lipton is the Edwin S. Lowe Professor of Neurology at the Albert Einstein College of Medicine in New York. He receives research support from the NIH: 2PO1 AG003949 (mPI), 1RF1 AG057531 (Site PI), 1UG3FD006795 (mPI), 1U24NS113847 (Investigator), U01 AT011005 (Investigator), 1R01 AG075758 (Investigator), 1R01 AG077639 (Investigator), 1RO1A1011875 (Investigator), 1RM1DA0055437 (Investigator), RO1AG080635 (Investigator), SG24988292 (Investigator), U19AGO76581 (Investigator), 1RO1NS123374 (Investigator), R61NS125153 (Investigator), and K23 NS107643 (Mentor). He also receives support from the Migraine Research Foundation and the National Headache Foundation and research grants from TEVA, Satsuma and Amgen. He serves on the editorial board of Neurology, senior advisor to Headache, and associate editor to Cephalalgia. He has reviewed for the NIA and NINDS, holds stock and stock options in Axon, Biohaven Holdings, CoolTech and Manistee; serves as consultant, advisory board member, or has received honoraria from: Abbvie (Allergan), American Academy of Neurology, American Headache Society, Amgen, Avanir, Axon, Axsome, Biohaven, Biovision, Boston Scientific, Dr. Reddy's (Promius), Electrocore, Eli Lilly, eNeura Therapeutics, Equinox, GlaxoSmithKline, Grifols, Lundbeck (Alder), Manistee, Merck, Pernix, Pfizer, Satsuma, Supernus, Teva, Trigemina, Vector, Vedanta. He receives royalties from Wolff's Headache 7th and 8th Edition, Oxford Press University, 2009, Wiley and Informa.; Dr. Ali Ezzati serves as consultant, advisory board member, or has received honoraria from: PCORI Health Care Horizon Scanning System, GlaxoSmithKline, Mist Research, and Corium. A.E is an Editorial Board Member of Journal of Alzheimer's Disease, A.E receives research support in part by grants from the National Institute of Health (NIA K23 AG063993, 2PO1 AG003949, AG080635), the Alzheimer's Association (SG-24-988292 ISAVRAD); Cure Alzheimer Fund, None of the sponsors had any role in the design, methods, data acquisition, analysis and preparation of the manuscript.;

### Author Declarations

Ethics committee/IRB of Eli Lilly Company and Alzheimer's Disease Neuroimaging Initiative gave ethical approval for this work

